# When Routine Errors Matter: The Effect of Non-Standardized Blood Pressure Measurement

**DOI:** 10.64898/2026.02.26.26347228

**Authors:** Paula Andrea Vesga-Reyes, Isabel Lucia Zapata-Vásquez, Diana Cristina Carrillo-Gómez, Juan Esteban Gómez-Mesa, Hoover Obdulio León-Giraldo, Carlos Enrique Vesga-Reyes

## Abstract

**Background:** Blood pressure (BP) is routinely measured during healthcare visits. A standardized measurement is essential to ensure accurate values, particularly in outpatient settings, where patient preparation, environment, and technique can significantly influence results.

**Methods:** A quasi-experimental study was conducted in adult outpatients. Demographic, anthropometric, and clinical data were collected through interviews and physical examination. BP was measured using a validated automated oscillometric device under four non-randomized predefined sequences. The standardized method followed international guideline recommendations, whereas the other three incorporated common errors observed in clinical practice (unsupported body position on the examination table, patient speaking, or legs crossed). Systolic and diastolic BP values were compared using the Friedman test and paired Wilcoxon tests with Holm adjustment. Effect sizes were expressed as median paired differences with interquartile ranges. Analyses were performed using R and Stata.

**Results:** A total of 295 participants were included (61% women; median age 56 years), with hypertension as the most frequent comorbidity (33%). Significant differences were observed across the four measurement models (p < 0.001). Compared with the standardized method, systolic BP was higher by +8 mmHg (M2), +2.5 mmHg (M3), and +4 mmHg (M4), while diastolic BP increased by +7 mmHg, +2 mmHg, and +2 mmHg, respectively. Clinically relevant differences (|Δ| ≥ 5 mmHg) occurred in up to 81% of systolic and 79% of diastolic measurements with M2.

**Conclusions:** Non-adherence to guideline-recommended BP measurement protocols leads to BP overestimation and misclassification of hypertension status, which may affect therapeutic decision-making and the use of pharmacological treatments.

## INTRODUCTION

Hypertension is one of the leading causes of death worldwide, contributing to approximately 10.4 million deaths annually (1,2). Accurate blood pressure (BP) measurement is essential for diagnosis and assessment (3). Improper measurement techniques or the use of inaccurate devices can lead to overestimation or underestimation of BP, potentially resulting in overdiagnosis and unnecessary treatment or, conversely, underdiagnosis and omission of treatment, both of which increase the risk of highly preventable cardiovascular diseases. For this reason, international guidelines have established standardized procedures for BP measurement, which include patient preparation, the environment, and the measurement technique itself (4,5).

Office blood pressure measurement correlates well with 24-hour ambulatory blood pressure monitoring. Therefore, accurate BP measurement in the clinical setting is critical for screening, diagnosis, and treatment (6). However, numerous factors are known to affect office BP readings, leading to variability in results ranging from 1 to 50 mmHg (7,8). The objective of this study was to determine whether there is a significant difference in BP values when following routine outpatient clinic procedures compared to standardized recommendations outlined in international guidelines.

## METHODS

### Data collection

A quasi-experimental study was conducted based on interviews and physical examinations during routine outpatient visits. Adult outpatients aged ≥18 years, with or without a prior diagnosis of hypertension, were eligible for inclusion. Pregnant women and individuals with anatomical abnormalities of the upper limbs were excluded. Eligible patients were informed about the study, and participation was voluntary. The study protocol was reviewed and approved by the institutional ethics committee. Given the minimal-risk nature of the study and the use of routine clinical procedures, the requirement for written informed consent was waived. Collected variables included age, sex, weight, height, body mass index (BMI), and pre-existing conditions such as type 2 diabetes mellitus, hypertension, chronic kidney disease, hypothyroidism, psychiatric disorders, as well as medications taken at the time of consultation, including antihypertensives, non-steroidal anti-inflammatory drugs (NSAIDs), antidepressants, erythropoietin, and corticosteroids.

### BP Measurement

Blood pressure was measured using a calibrated and validated blood pressure monitor manufactured by OMRON, model HEM-RML31. An appropriately sized cuff (22–42 cm upper arm circumference) was used. Blood pressure was measured by the study personnel (medical staff) using the same automatic blood pressure monitor. All patients were required to have an empty bladder, and it was verified that they had not engaged in physical activity or consumed tobacco, caffeine or energy drinks within 30 minutes prior to entering the consultation room and remain seated and relaxed for 3-5 minutes. Three initial readings were obtained, with at least one minute between each, and the average of the last two values was recorded. The arm with higher blood pressure reading was selected.

Each patient underwent blood pressure measurement using four different sequencing models according to enrollment (Figure 1). The standard measurement method (M1) was the one that adhered to all recommendations from international guidelines. In this method, it was ensured that the patient did not speak, had their arm supported at heart level, the cuff placed on a bare arm, feet flat on the floor without crossing the legs, and back supported against the chair. The other three methods included common errors observed in routine outpatient practice. The second method, unsupported position method (M2), involved the patient not having their feet, arm, or back supported, in the examination table. The third method, talking method (M3), followed the standard procedure; however, the patient spoke during the measurement. The fourth method, the crossed-legs method (M4), also followed the standard procedure, but the patient had their legs crossed.

**Figure 1.**
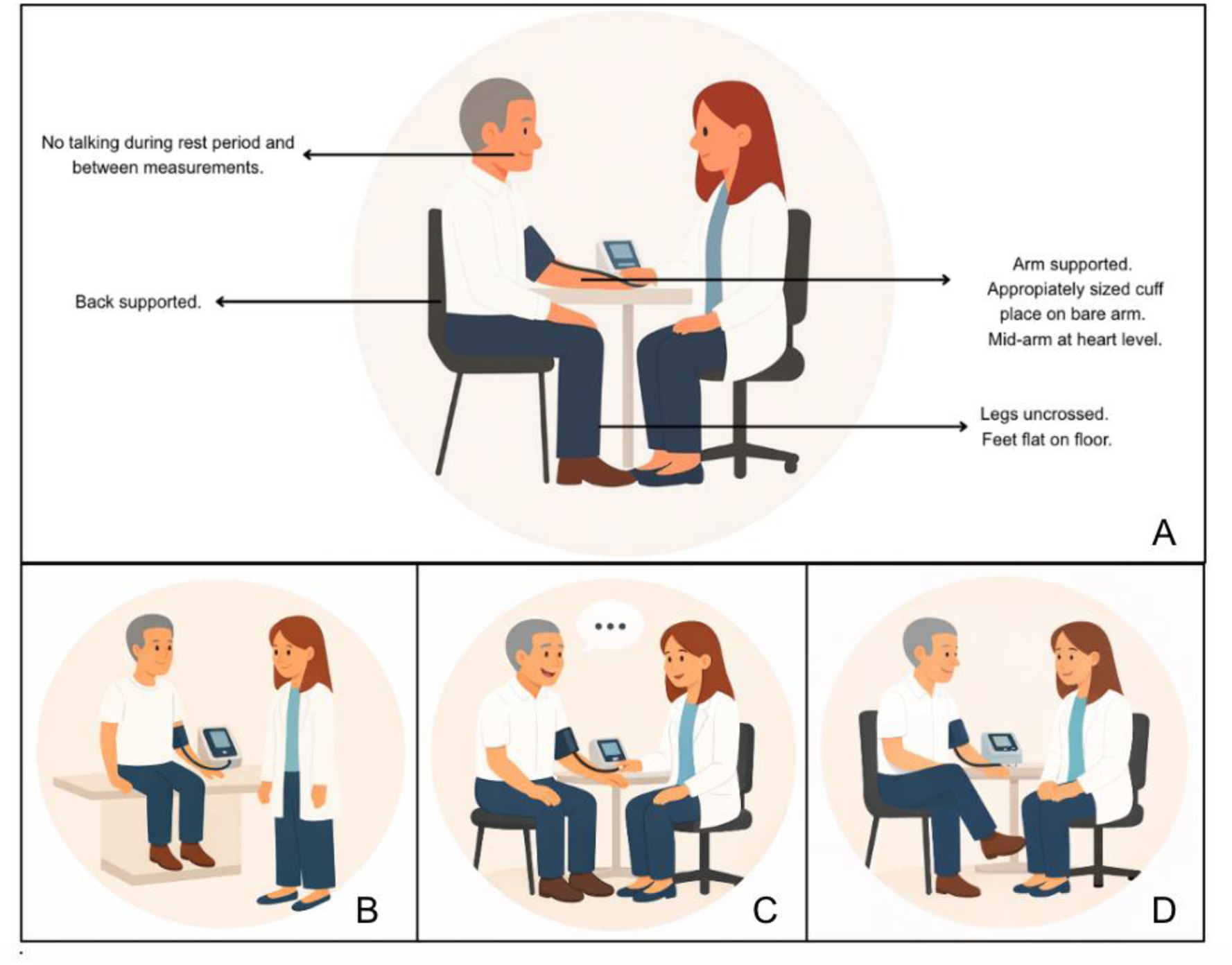
A) Standard measurement method (M1). B) Unsupported position method (M2). C)Talking method (M3) D) Crossed-legs method (M4)

### Outcomes

The primary outcome was the difference in systolic, diastolic, and mean blood pressure between each measurement model and the standardized method (M1). For pairwise analyses we computed paired differences Δ = Mi − M1 (i = 2, 3, 4). Clinical relevance was prespecified as |Δ| ± ≥ 5 mmHg; additional thresholds (≥10, ≥15, ≥20 mmHg) were explored descriptively.

### Statistical analysis

Baseline characteristics were summarized using medians and interquartile ranges (IQRs) for continuous variables (distributions assessed with Shapiro–Wilk) except for height, which was summarized using the mean and standard deviation. Categorical variables were summarized as absolute and relative frequencies. Potential outliers were visually assessed using boxplots and were not removed automatically.

Overall differences among the four methods were evaluated using the Friedman test (non-parametric repeated-measures ANOVA), a non-parametric alternative to repeated-measures ANOVA, suitable when the assumption of normality is violated, as in this study. Following the global Friedman test, pairwise comparisons against the standard model (M1) were performed using the two-sided Wilcoxon signed-rank test. The p-values were adjusted for multiplicity using Holm’s method within each family of comparisons (M2–M1, M3–M1, M4–M1) for each measurement (SBP, DBP). The effect size was reported as the median of the paired differences (Δ), along with the IQR and adjusted p-values (Hodges–Lehmann estimate) (9).

For result presentation, box-and-whisker plots were used along with waterfall plots, that rank individual Δ values (Mi vs M1) from smallest to largest. Positive values (>0) indicate that the alternate model (Mi) yielded higher readings than the standard (M1). The green line (0 mmHg) represents no bias; red dashed lines (±5 mmHg) indicate the threshold for clinical relevance. Subgroup tables (hypertension history, sex, and age ≥65 years) were produced to contextualize inter-individual variability. All data processing and statistical analyses were conducted using R software (version 4.4.1) and Stata 15.0.

## RESULTS

### Characterization of the study population

The analyzed cohort included 295 participants, the majority of whom were female (61%) with a median age of 56 years (IQR: 33–70). Regarding nutritional status, 44.8% had a normal body mass index (BMI), whereas 52.9% were overweight (39.7%) or obese (13.2%). Among comorbidities, arterial hypertension was the most prevalent (33%), followed by dyslipidemia (23%) and primary hypothyroidism (14%) (Table 1). The most frequently prescribed drug class was angiotensin II receptor blockers (ARBs) (25%), followed by beta-blockers (22%), diuretics (14%), and calcium channel blockers (CCBs) (13%). Additional medications that could potentially influence blood pressure values were considered, including non-steroidal anti-inflammatory drugs (NSAIDs), opioids, psychiatric treatments, erythropoietin, and SGLT2 and GLP1 inhibitors (Table 1).

**Table 1.**
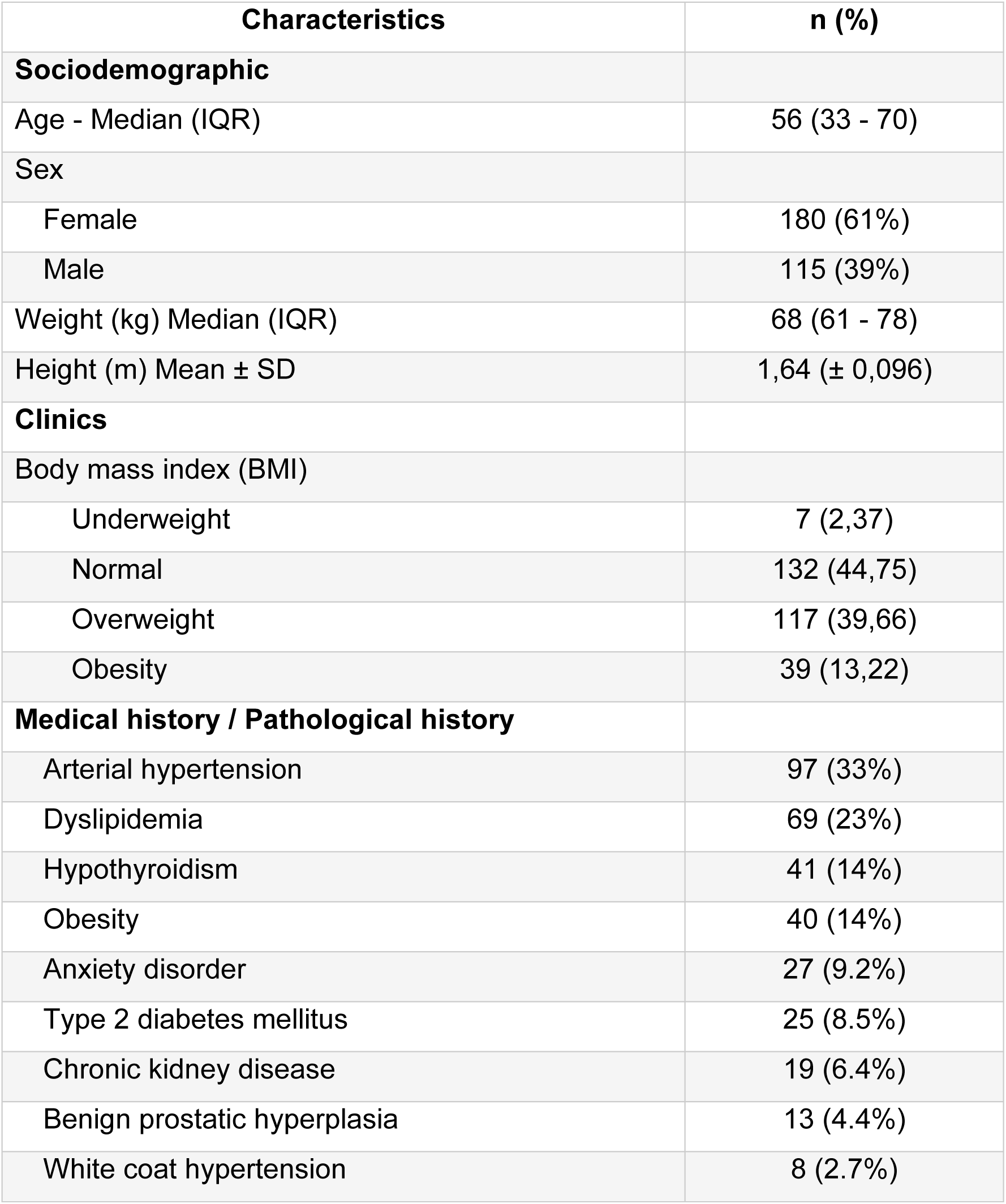
Sociodemographic and clinical characteristics of the study population.

When comparing overall SBP and DBP values across the four models, statistically significant differences were observed according to Friedman’s test (p < 0.001) (Figures 2). Box plots display SBP (top) and DBP (bottom) values according to measurement-sequence position within the predefined measurement-sequence models. SBP1–SBP4 and DBP1–DBP4 denote the first through fourth sequential blood pressure measurements obtained during the study protocol. Boxes represent interquartile ranges, center lines indicate medians, whiskers show dispersion, and points represent outliers.

**Figure 2.**
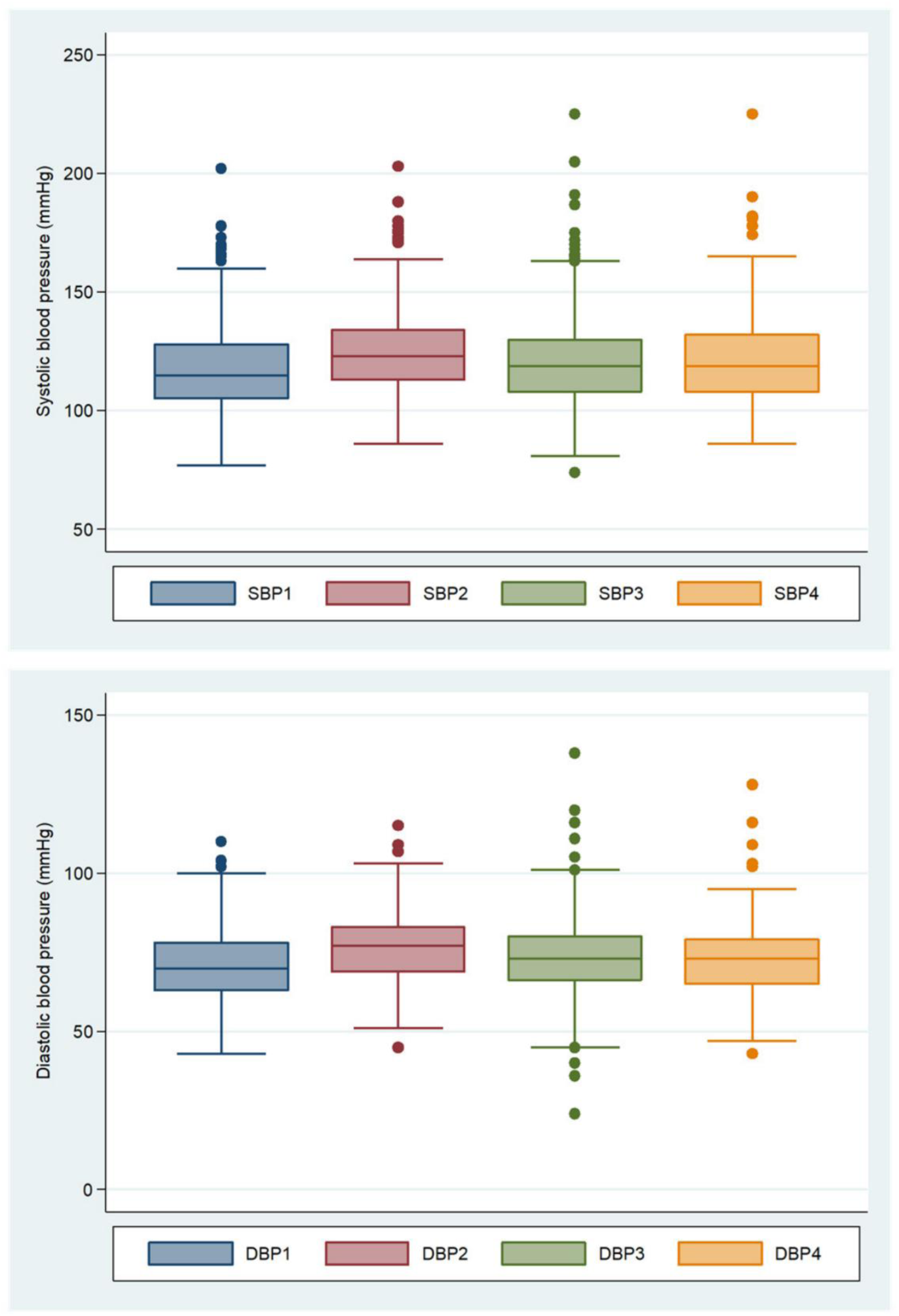
Distribution of systolic blood pressure (SBP) and diastolic blood pressure (DBP) by measurement-sequence position

### Comparison of systolic and diastolic blood pressure values by blood pressure measurement model

Figures 3 illustrate within-patient differences between each alternative model (M2, M3, M4) and the standard model (M1) for SBP and DBP. Positive values indicate that the alternative models yielded higher readings than the standard model. The green line at zero represents absence of bias, while the dashed red lines at ±5 mmHg denote the threshold of clinical relevance. For SBP, median differences (Mi−M1) were +8 mmHg for M2, +2.5 mmHg for M3, and +4 mmHg for M4, all statistically significant (p < 0.001). For DBP, similar findings were observed, M2 showed a median difference of +7 mmHg, whereas M3 and M4 showed +2 mmHg each, also significant (p < 0.001).

**Figure 3.**
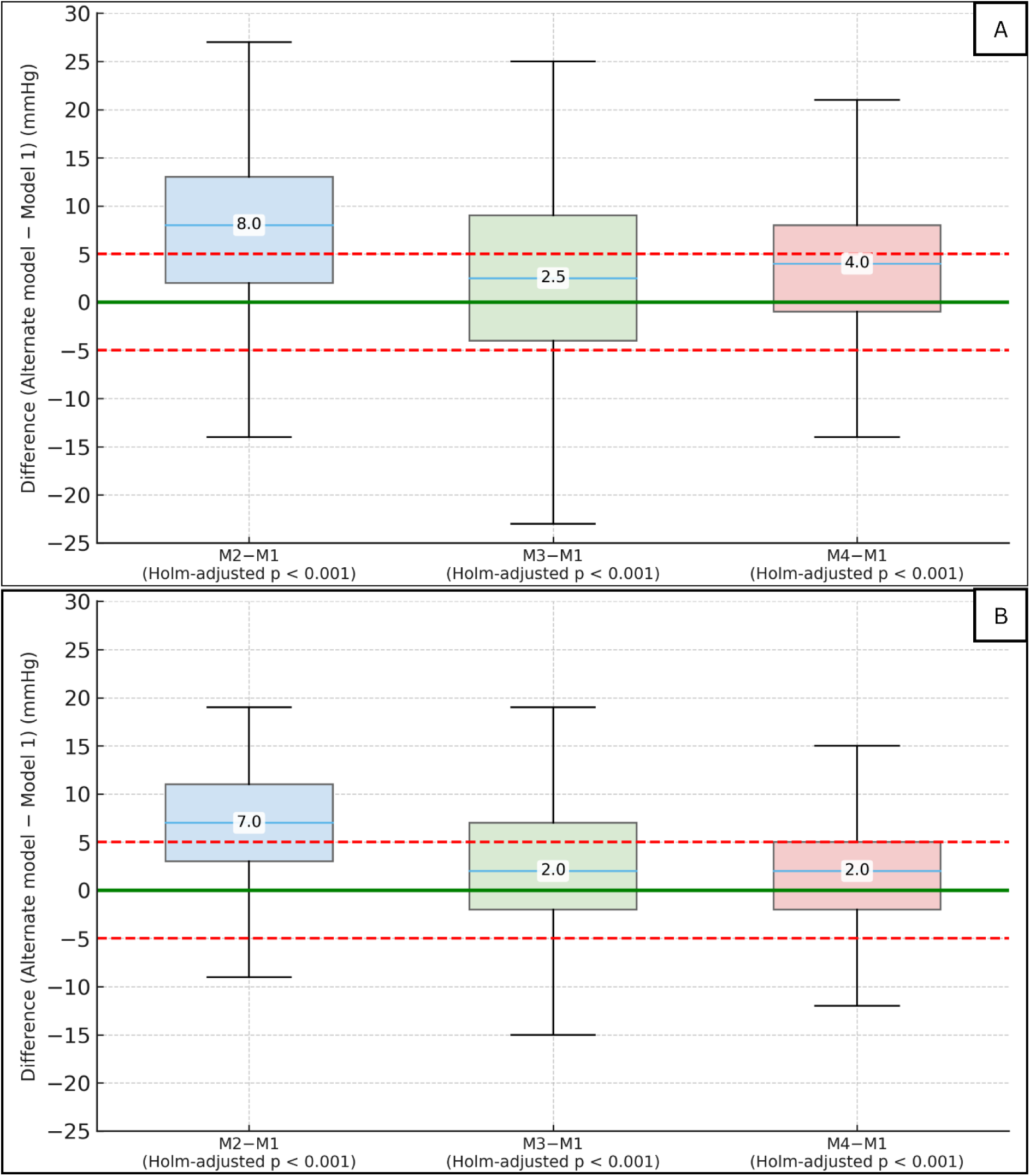
Comparison of systolic blood pressure (A) and diastolic blood pressure (B) with the standard model

The paired differences (Δ = Mi − M1) were predominantly positive. Relative to the standard model SBP exceeded the +5 mmHg threshold in 81% of cases with M2, 66.7% with M3, and 60.3% with M4. For DBP, a similar pattern was noted, with positive differences exceeding the +5 mmHg threshold in 79%, 50.2%, and 43.1% of cases, respectively (Figures 3 and 4). We also quantified the proportion of patients whose SBP and DBP differences crossed more stringent thresholds (+10, +15, +20 mmHg), The following figures summarizes, for SBP and DBP, the proportion of participants whose absolute difference from the standard model (M1) exceeds the thresholds of clinical relevance (|Δ| ≥ 5 mmHg). As expected, increasing the threshold (≥10, ≥15, and ≥20 mmHg) reduces the proportion of patients exceeding it. Across all thresholds, M2 shows the highest proportions, M3 an intermediate position, and M4 the lowest; the only exception occurs in DBP at the ≥20 mmHg threshold, where M3 exceeds M2 (3.7% vs. 1.4%, respectively). (Figure 4).

**Figure 4.**
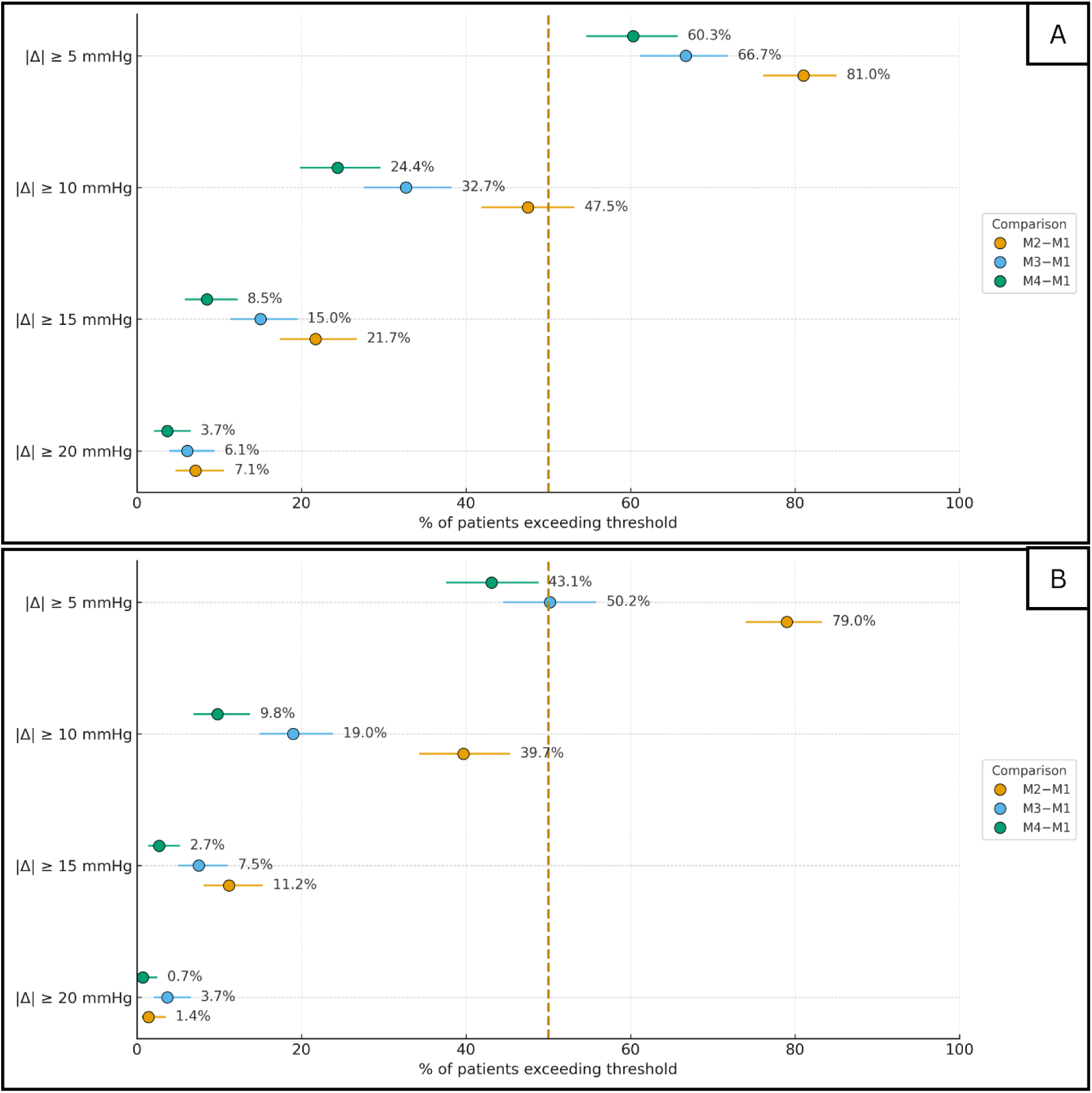
Proportion of patients with clinical relevance systolic blood pressure (A) and diastolic blood pressure (B) measurement

Subgroup analyses of SBP and DBP were performed according to hypertension status, sex, and age ≥65 years (Table 2). Compared with the standard method (M1), the unsupported position method (M2) showed a consistent and strong positive bias across all subgroups, with widespread clinical relevance. In contrast, the talking method (M3) demonstrated a moderate positive bias with heterogeneity among subgroups. Significant differences were observed in patients without hypertension, in both sexes, and in younger adults, but not in those with a history of hypertension (p=0.845) or in older adults (p=0.245). The crossed-legs method (M4) showed a positive bias of small-to-moderate magnitude, slightly higher in men (median, +5 mmHg), with all estimated differences reaching statistical significance (p<0.001).

**Table 2.**
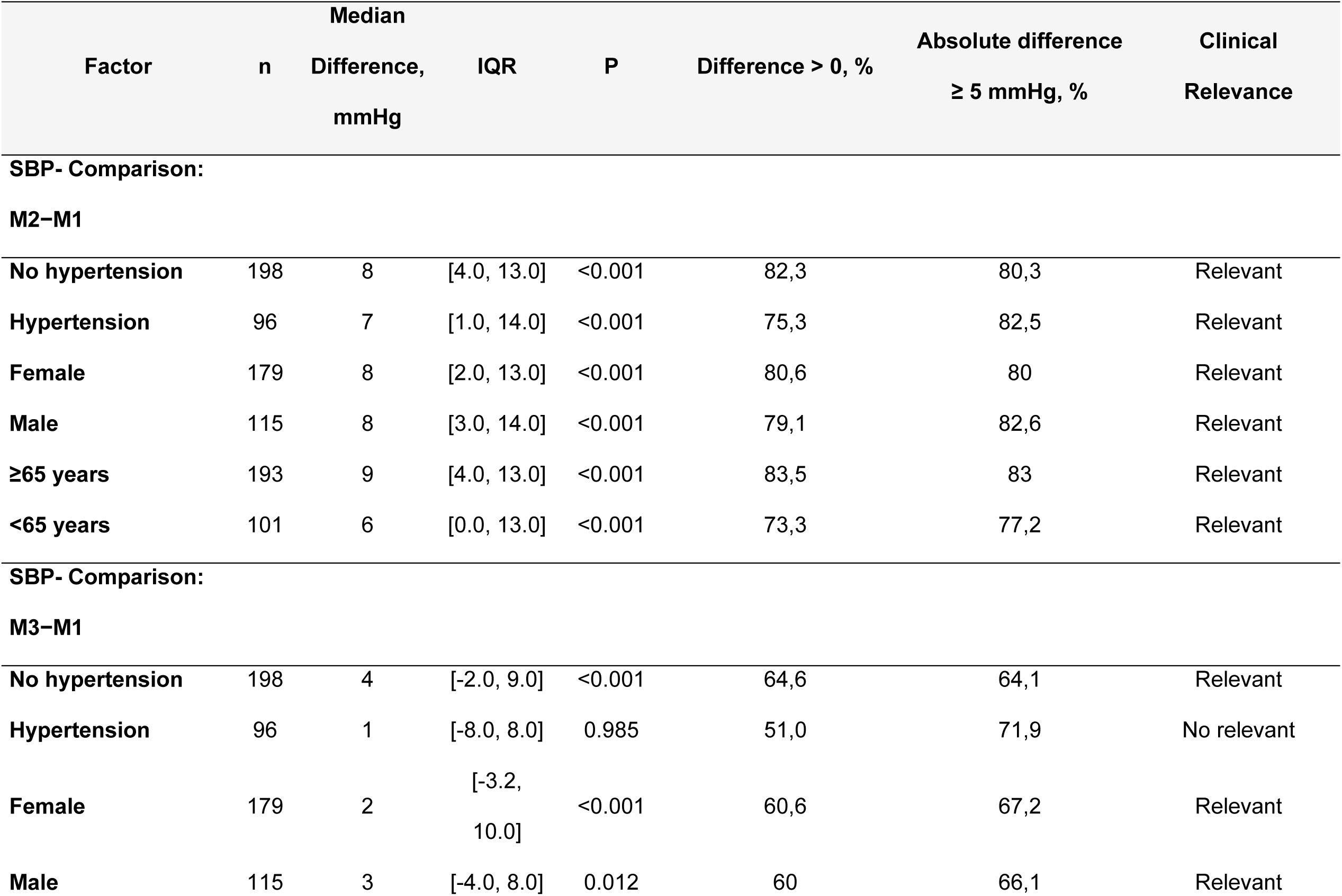
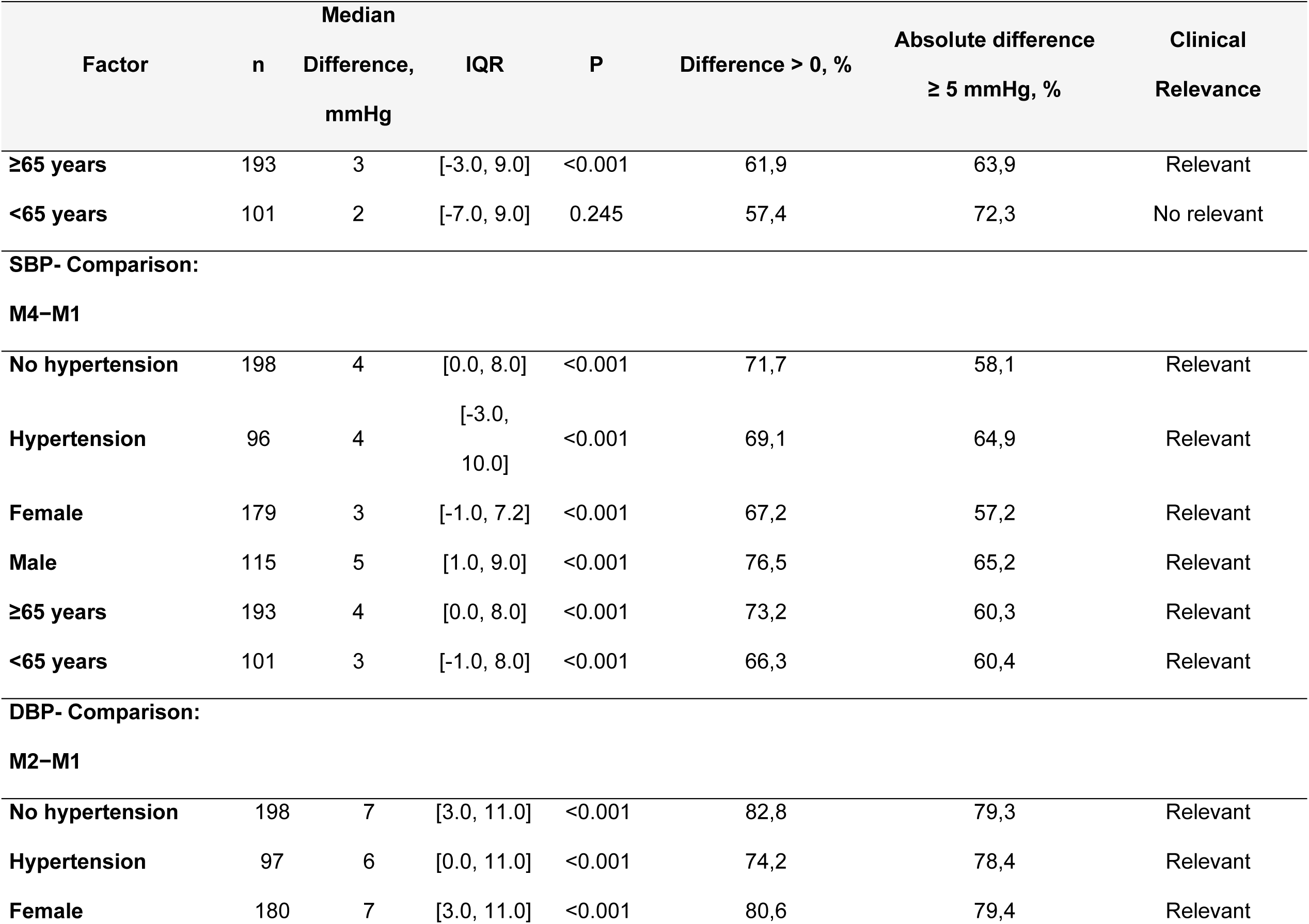
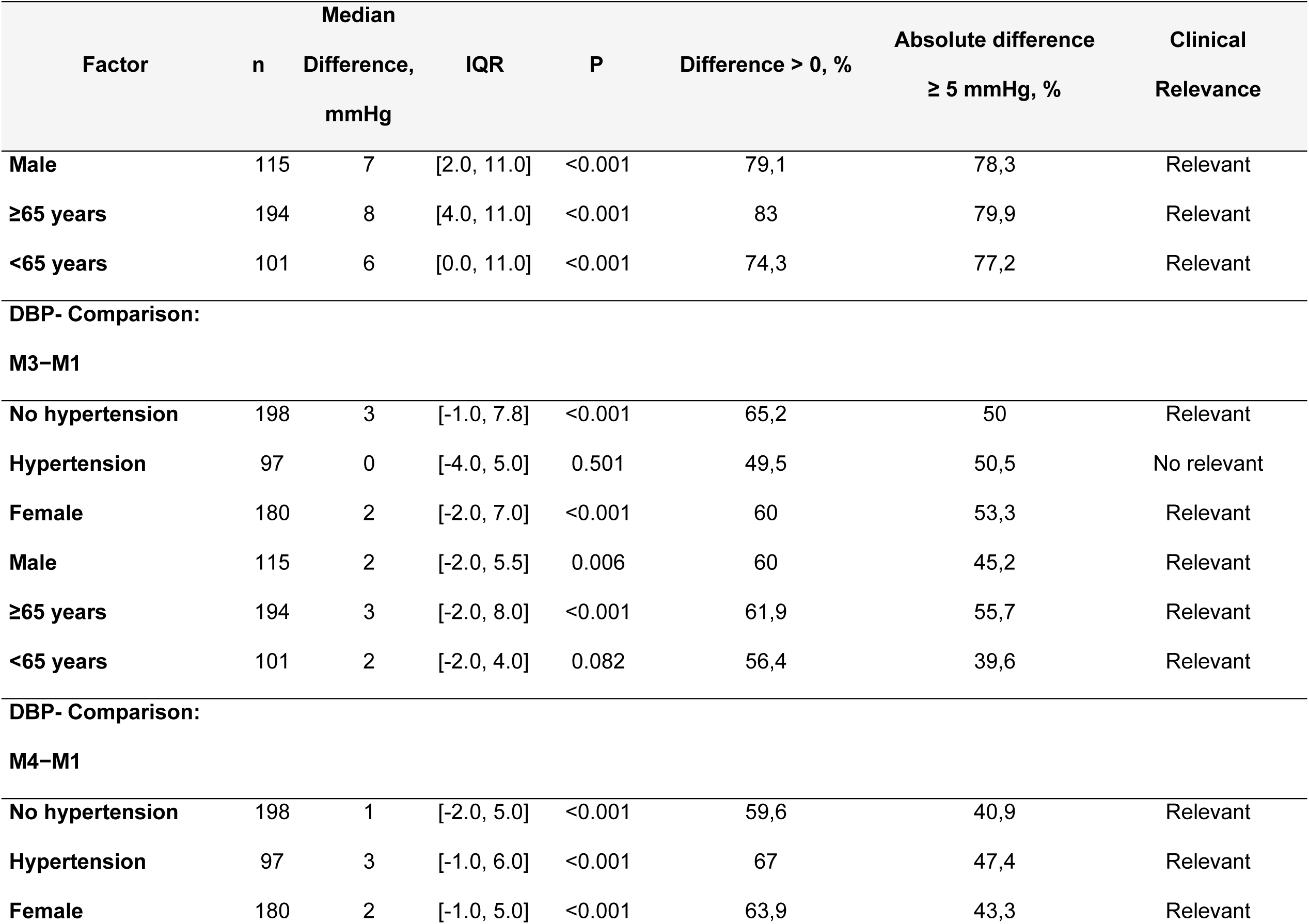
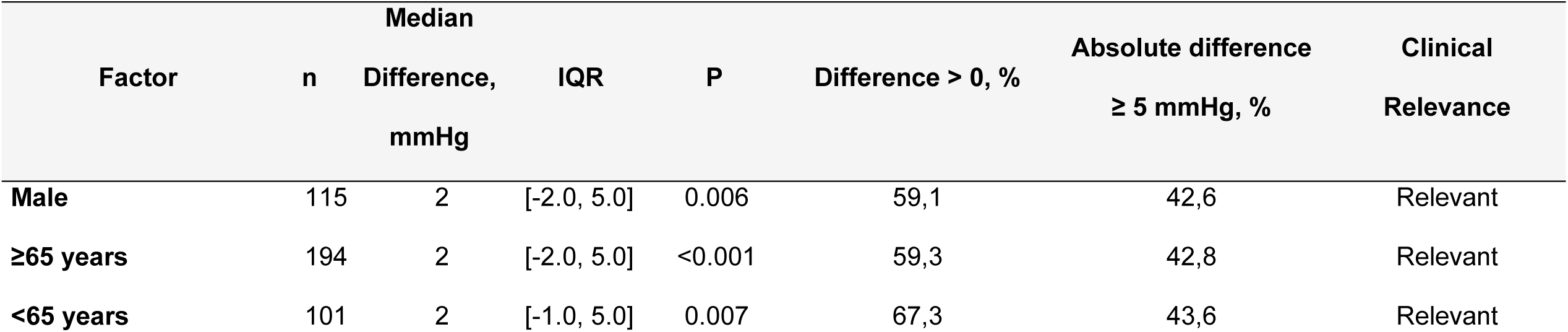
Differences in systolic (SBP) and diastolic (DBP) blood pressure between the alternate models and the standard model across subgroups.

Similarly, when comparing DBP values, M2 showed a strong and consistent positive bias across all subgroups, with all comparisons reaching statistical significance (p < 0.001). M3 demonstrated a small-to-moderate positive bias with subgroup heterogeneity; differences were not significant in patients with hypertension (p = 0.501) or in older adults (p = 0.082). For M4, the bias was positive, with small-to-moderate magnitude, slightly higher in patients with hypertension (median +3 mmHg).

## DISCUSSION

In this study, we evaluated the agreement between several blood pressure (BP) measurement models and a standard reference model in a cohort of nearly 300 outpatients, identifying significant differences among them. Our findings highlighted that alternative models incorporating common procedural errors, such as lack of back or arm support, patient talking during measurement, or sitting with crossed legs, systematically overestimated both systolic and diastolic blood pressure compared with the standardized method. Notably, the magnitude of this bias exceeded clinically relevant thresholds (+5 mmHg) in a substantial proportion of patients, particularly in the unsupported position method (M2), which represented the model with the greatest number of errors. The consistent positive bias observed in M2 across all subgroups suggests a systematic overestimation, which is worrisome, given that such discrepancies may lead to misclassification of hypertension status and unnecessary initiation or intensification of antihypertensive therapy. Although smaller, the biases observed in M3 and M4 remain clinically relevant, reinforcing that even seemingly minor deviations from standardized technique can meaningfully influence BP values.

Our results align with previous research demonstrating that variability in measurement technique significantly impacts BP accuracy and clinical decision-making (10,11). Hypertension guidelines emphasize accurate BP measurement as the cornerstone for hypertension diagnosis and management, yet, in the daily practice, deviations from standardized BP measurement protocols, even those that may appear minor or are commonly observed, significantly overestimate both SBP and DBP (12,13). In our study, more than 60% of patients exceeded the +5 mmHg threshold for SBP in M2 and nearly 80% for DBP, underscoring the potential clinical consequences. Our findings demonstrate that frequent procedural errors during outpatient BP assessment, including inadequate support of the back or arm, patient conversation during the reading, or sitting with crossed legs, consistently result in an upward bias of both systolic and diastolic values. Correct BP measurement is a critical step in the diagnosis and management of hypertension; however, in routine clinical practice many of the recommended procedures are often overlooked (14). In busy primary care settings, patients may remain seated in waiting rooms for extended periods before measurement, and assessments are frequently performed without full adherence to established protocols (15–17).

Accurate BP measurement remains challenging, as multiple procedural and physiological factors contribute to variability (18). Our findings demonstrate clinically meaningful differences in blood pressure values obtained during usual care compared with those measured under standardized conditions recommended. This underscores that proper adherence to measurement techniques is essential not only for minimizing the white coat effect but also for ensuring accurate classification of blood pressure control. These discrepancies between usual practice and guideline-based measurement, have significant clinical implications, including the potential overdiagnosis of hypertension, inappropriate treatment intensification, and unnecessary patient exposure to pharmacological therapies with associated risks (17,19).

These findings support previous studies showing that systematic errors, even of small magnitude, have significant implications for hypertension diagnosis. Prior modeling studies have shown that a +3–5 mmHg bias in SBP may increase hypertension prevalence estimates by up to 43%, while a +5 mmHg bias in DBP could more than double the number of diagnosed cases (20). Similarly, modeling studies also indicate that a +10 mmHg overestimation may raise hypertension prevalence by 50–63%, overtreatment of approximately 3.5 million patients (21). Furthermore, pseudoresistance attributed to measurement error has been reported in as many as 33% of patients with apparent uncontrolled resistant hypertension

(22). These data collectively reinforce that procedural errors are not merely nuisances but can fundamentally alter patient trajectories and health system performance.

The literature also shows that measurement errors are not only uncommon but often cumulative factors such as arm position, anxiety, poor technique, and incorrect cuff size, all introduce significant variability (23). Automated office BP, when performed under standardized conditions (patient resting, alone), has been shown to correlate more strongly with daytime ambulatory BP and to mitigate the white-coat effect, thus providing more reliable estimates than manual methods (24). From a public health perspective, inaccurate measurement also inflates population-level estimates of hypertension prevalence, misguides resource allocation, and compromises the validity of quality indicators used in health systems (25).

Our study has several strengths, including a relatively large sample size, the use of a validated device, and rigorous application of both standardized and error-prone measurement conditions. In addition, subgroup analyses across sex, age, and hypertension status enhance the external validity of our findings. However, limitations must be acknowledged. First, the cross-sectional design precludes assessment of longitudinal outcomes related to misclassification. Second, our results may not be generalizable to populations with different demographic or clinical profiles, or to other devices not included in this study. Third, no randomization was performed. Instead, four predefined measurement-sequence models were implemented and assigned sequentially according to the order of study enrollment to avoid bias arising from the possibility that the first blood pressure measurement would be systematically higher because of the measurement-order effect.

## PERSPECTIVES

This study provides robust and quantifiable evidence that common errors during outpatient BP measurement result in systematic overestimation of both SBP and DBP, with a substantial proportion of patients exceeding clinically meaningful thresholds. These findings underscore the need for rigorous adherence to standardized protocols in clinical practice, highlight the risks of diagnostic misclassification, and call for renewed efforts to implement feasible strategies that improve BP measurement quality in routine care. Future research should focus on the longitudinal consequences of these errors, their impact on treatment patterns, and the development of interventions to enhance standardization in real-world clinical workflows.

## NOVELTY AND RELEVANCE

### What Is New?

This study provides direct evidence that common deviations from standardized BP measurement protocols in outpatient practice systematically overestimate both systolic and diastolic blood pressure, with differences frequently exceeding clinically relevant thresholds.

### What Is Relevant?

Our findings highlight that even small procedural errors, such as lack of back or arm support, speaking during the measurement, or leg crossing, significantly alter blood pressure values. These inaccuracies can influence hypertension diagnosis, assessment of blood pressure control, and therapeutic decisions.

### Clinical/Pathophysiological Implications?

These data underscore the critical importance of adhering to international guidelines for BP measurement in clinical practice. Failure to follow standardized protocols can lead to overdiagnosis, unnecessary initiation or escalation of antihypertensive therapy, and increased exposure to drug-related adverse effects, ultimately compromising patient safety and cardiovascular risk management.

## Data Availability

The data presented in this study are included in the article. Deidentified participant-level data underlying the reported results, the data dictionary, and the analytic code can be made available by the corresponding author upon reasonable request, subject to institutional approval and execution of a data use agreement, in accordance with applicable ethical and privacy regulations

## DISCLOSURES

There are no conflicts of interest.

Artificial intelligence–based tools were used to assist with language editing and stylistic refinement. The authors take full responsibility for the content, interpretation, and conclusions of this manuscript.

